# Huge Excess Mortality due to the delta strain of COVID-19 in Japan in August and September, 2021

**DOI:** 10.1101/2020.07.09.20143164

**Authors:** Junko Kurita, Tamie Sugawara, Yasushi Ohkusa

## Abstract

**Background:** No remarkable mortality attributable to COVID-19 confirmed by PCR test has been observed in Japan.

**Object:** We sought to quantify excess mortality using the National Institute of Infectious Diseases (NIID) model.

**Method:** We applied the NIID model to deaths of all causes from 1987 up through October, 2021 for the whole of Japan and up through August, 2021 for Tokyo.

**Results:** Results in Japan show huge number of excess mortality, up to 10 thousands in the two months, in August and September, 2021. On the other hand, in Tokyo, 1323 excess mortality was detected

**Discussion and Conclusion:** We detected substantial excess mortality in Japan in August and September, 2021 and in Tokyo in August, 2021. It might be important to continue to monitor excess mortality of COVID-19 carefully hereafter.

## 1. Introduction

To date, excess mortality has mainly been used to assess the social effects of influenza activity [1–6]. However, since the emergence of COVID-19, excess mortality attributable to COVID-19 has been attracting attention [7] as a measure of the total effects of the disease because it can reflect cases which have not been identified as polymerase chain reaction (PCR) positive. Especially in Japan, PCR tests administered per capita have been few. Therefore, concern has arisen about the possibility that some deaths caused by COVID-19 have not been recognized heretofore. Moreover, excess mortality related to COVID-19 might be expected to contribute to evaluation of vaccine effects. For these evaluations, the estimated excess mortality without the effects of a vaccine should be regarded as a baseline. Nevertheless, no such a trial has been undertaken to date. This study might be the first trial to measure that figure in Japan.

As of the end of 2021, the COVID-19 outbreak showed about 1.73 million patients and about 1.84 thousand deaths from the outbreak have been reported in Japan. Although Japan has about one third of the population of the U.S., these figures are vastly different in scale from those of the U.S., which has reported 54.8 million cases of morbidity and 826 thousand deaths [8]. In light of the much lower number of patients in Japan, some criticism has arisen that low PCR testing rates might have led to the lower number of documented patients [9]. In this sense, one might regard the number of deaths as reflecting the actual situation in Japan, but with no testing-related bias.

Concerning deaths, the case-fatality rate (CFR) is about 5%. In fact, the CFRs in both countries are not much different. The lower PCR testing in Japan might be related to some problems. Therefore, we specifically examined excess mortality attributable to COVID-19 in Japan, irrespective of the cause of death.

In Japan, excess mortality was estimated using the National Institute of Infectious Diseases (NIID) model [10], which has been the official procedure for more than ten years. It was applied to two data sources: the national monthly deaths of all causes and the respective weekly pneumonia and influenza deaths in the 21 largest cities and their total. The latter is published regularly in Japanese during the influenza season as https://www.niid.go.jp/niid/ja/flu-m/2112-idsc/jinsoku/131-flu-jinsoku.html. Unfortunately, that publication ceased in March 2020 because it is intended for influenza. The first peak in Japan was April 3, 2020[11]: excess mortality cannot be detected until March. Instead, we applied NIID model to the all causes of death in the whole of Japan to evaluate impact of the outbreak of COVID-19.

## 2. Method

Excess mortality is defined as the difference between the actual number of deaths and an epidemiological threshold. The epidemiological threshold is defined as the upper bound of the 95% confidence interval (CI) of the baseline. The baseline is defined as the number of deaths that are likely to have occurred if an influenza outbreak had not occurred. Therefore, if the actual deaths are fewer than the epidemiological threshold, then excess mortality is not inferred.

The data used for this study were monthly deaths of all causes from 1987 through October 2021 for the whole of Japan and August 2021 for Tokyo [12]. NIID model, the Stochastic Frontier Estimation [13–19], is presented as

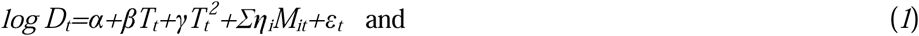

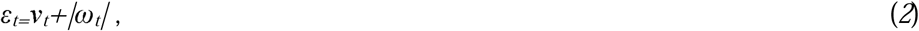

where *D*_*t*_ represents all causes of death in month/year *t, T*_*t*_ denotes the linear time trend, and *M*_*it*_ is the dummy variable for a month, which is one if *t* is the *i-*th month and otherwise zero. Moreover, *v*_*t*_ and *ω*_*t*_ are stochastic variables as *v*_*t*_ *∼N*(*0, μ*^2^) and *ω*_*t*_ *∼N*(*0,ξ*^2^); they are mutually independent. Although *v*_*t*_ represents stochastic disturbances, *ω*_*t*_ denotes non-negative deaths attributable to influenza. These disturbance terms in this model are parameterized by two parameters: *ξ*/*μ* and (*μ*^*2*^*+ξ*^*2*^)^*0*.*5*^. If the null hypothesis *ξ*/*μ*=0 is not rejected, then the Stochastic Frontier Estimation model is inappropriate.

Study areas were the whole of Japan and its capital, Tokyo. Study period foe estimation was from 1987 to October 2021 for the whole of Japan and up through August, 2021 for Tokyo. We adopted 5% as significant level.

## 3. Results

Table 1 summarized the estimation results in the whole of Japan and Table 2 for Tokyo. Figure 1 presents observed deaths, the estimated baseline, and its threshold in Japan. Figure 2 specifically depicts the last year in Japan. We found 12 and 104 excess mortality in August and October, 2020, and 260, 165 and 584 in May, June and October, 2021. These were 0.0, 0.1, 0.2, 0.1 and 0.5% of the baseline. Moreover, we found huge number of excess mortality in August and September, 2021, which were estimated as 4106 and 5854, and were 3.8% and 5.5 % of the baseline.

**Table 1.** NIID Model estimation results since 1987 until October 2021 in Japan

| Explanatory variables | Estimated coefficients | *p*-value |
| --- | --- | --- |
| Constant | 11.12 | <0.0004 |
| Time trend | 0.001537 | <0.0004 |
| Time trend <sup>2</sup> | 0.0000009763 | 0.423 |
| January | 0.07075 | <0.0004 |
| February | -0.05631 | <0.0004 |
| March | -0.01671 | 0.053 |
| April | -0.1027 | <0.0004 |
| May | -0.1247 | <0.0004 |
| June | -0.2126 | <0.0004 |
| July | -0.1776 | <0.0004 |
| August | -0.1710 | <0.0004 |
| September | -0.2083 | <0.0004 |
| October | -0.1197 | <0.0004 |
| November | -0.08768 | <0.0004 |
| $\xi/\mu$ | 2.386 | <0.0004 |
| $(\mu^2 + \xi^2)^{0.5}$ | 0.04934 | <0.0004 |

Note: The number of observations was 414. The estimated log likelihood was 915.5. $\xi^2$ denotes the variance of the non-negative disturbance term. $\mu^2$ is the variance of the disturbance term.

**Table 2.**
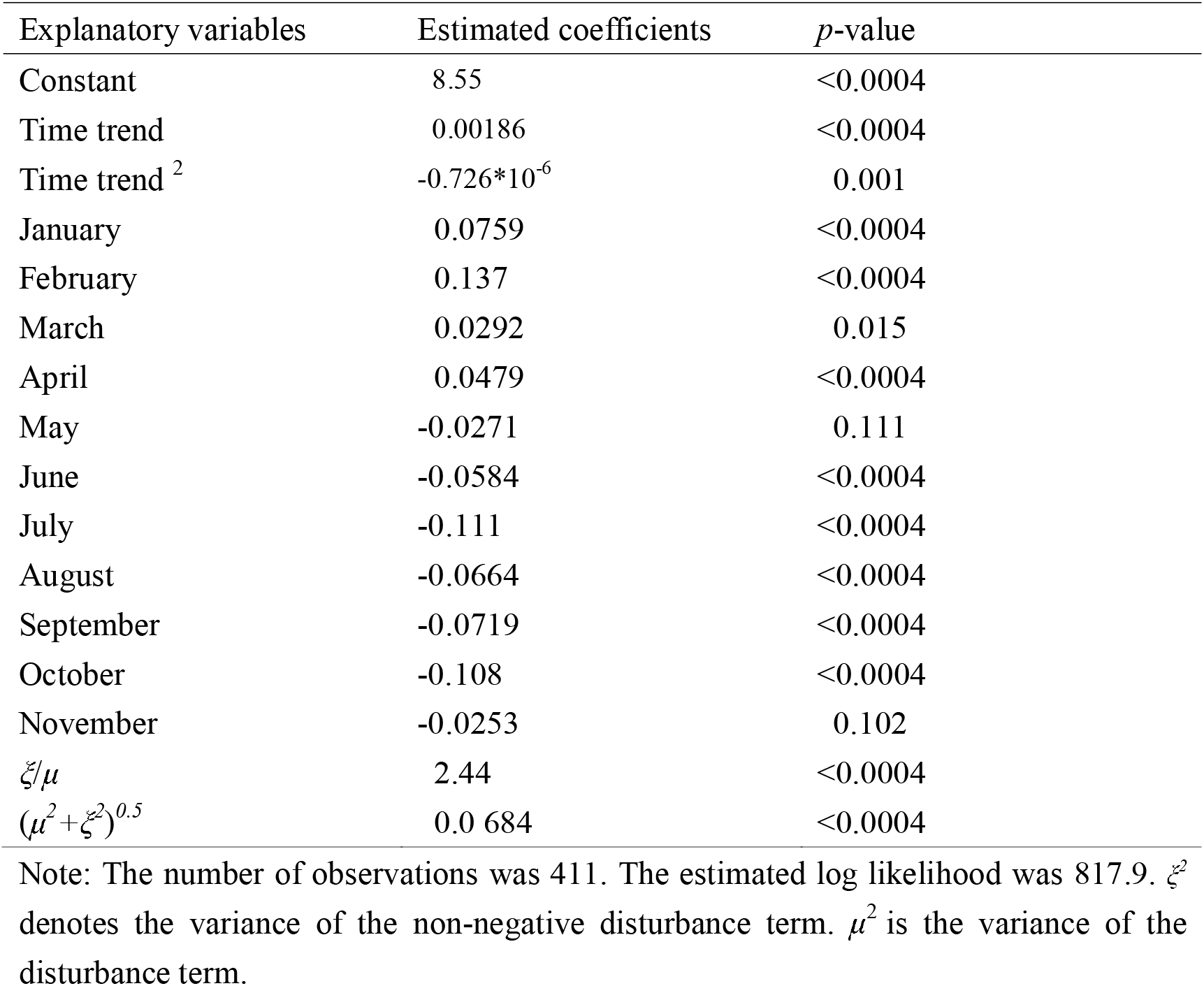
NIID Model estimation results since 1987 until August 2021 in Tokyo

| Explanatory variables | Estimated coefficients | *p*-value |
| --- | --- | --- |
| Constant | 8.55 | <0.0004 |
| Time trend | 0.00186 | <0.0004 |
| Time trend <sup>2</sup> | -0.726*10<sup>-6</sup> | 0.001 |
| January | 0.0759 | <0.0004 |
| February | 0.137 | <0.0004 |
| March | 0.0292 | 0.015 |
| April | 0.0479 | <0.0004 |
| May | -0.0271 | 0.111 |
| June | -0.0584 | <0.0004 |
| July | -0.111 | <0.0004 |
| August | -0.0664 | <0.0004 |
| September | -0.0719 | <0.0004 |
| October | -0.108 | <0.0004 |
| November | -0.0253 | 0.102 |
| $\xi/\mu$ | 2.44 | <0.0004 |
| $(\mu^2 + \xi^2)^{0.5}$ | 0.0 684 | <0.0004 |

Note: The number of observations was 411. The estimated log likelihood was 817.9. $\xi^2$ denotes the variance of the non-negative disturbance term. $\mu^2$ is the variance of the disturbance term.

**Figure 1:**
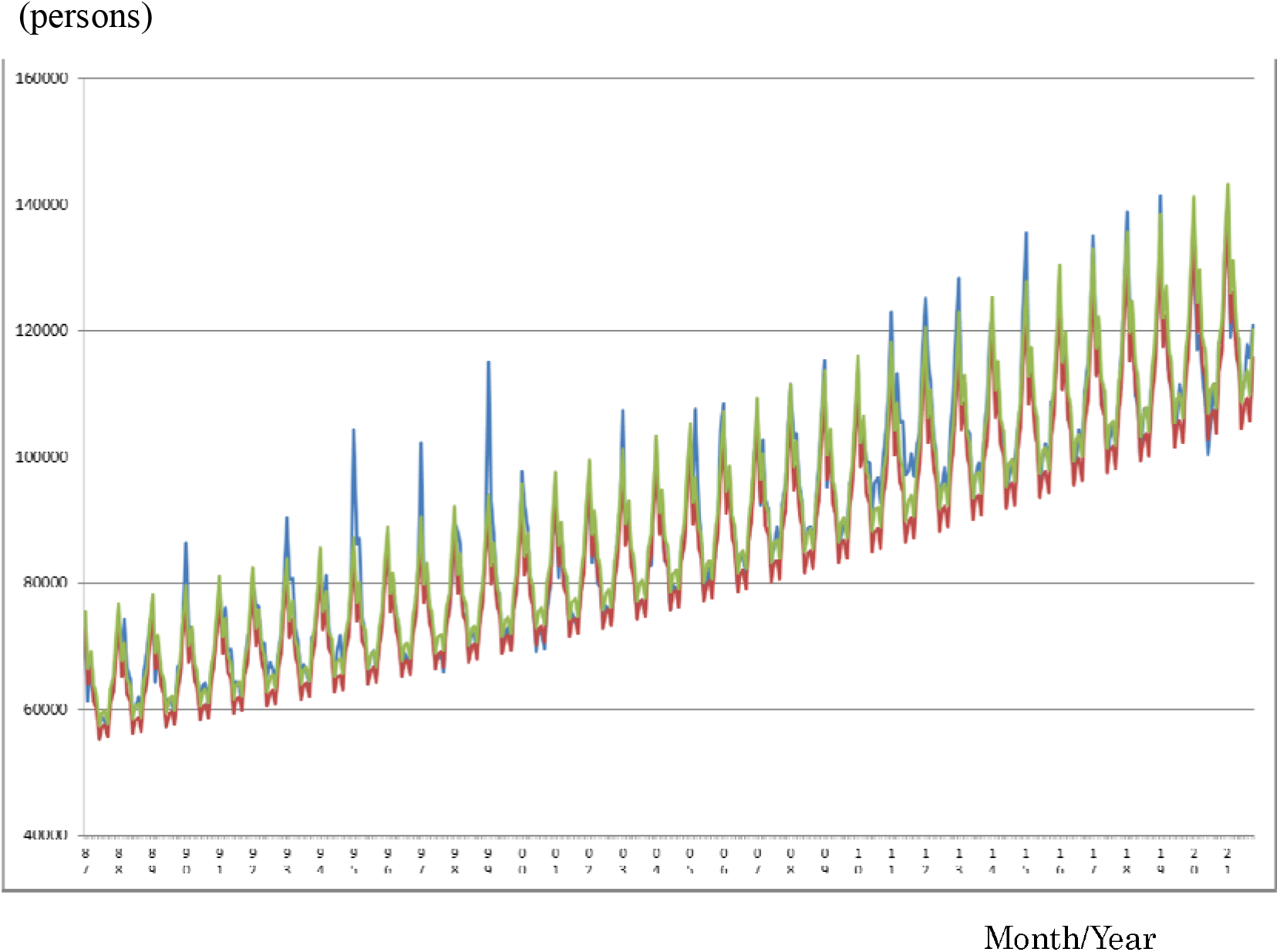
Observations of the estimated baseline and threshold since 1987 until October 2021 in Japan Note: The blue line represents observations. The red line represents the estimated baseline. The green line shows its threshold.

**Figure 2:**
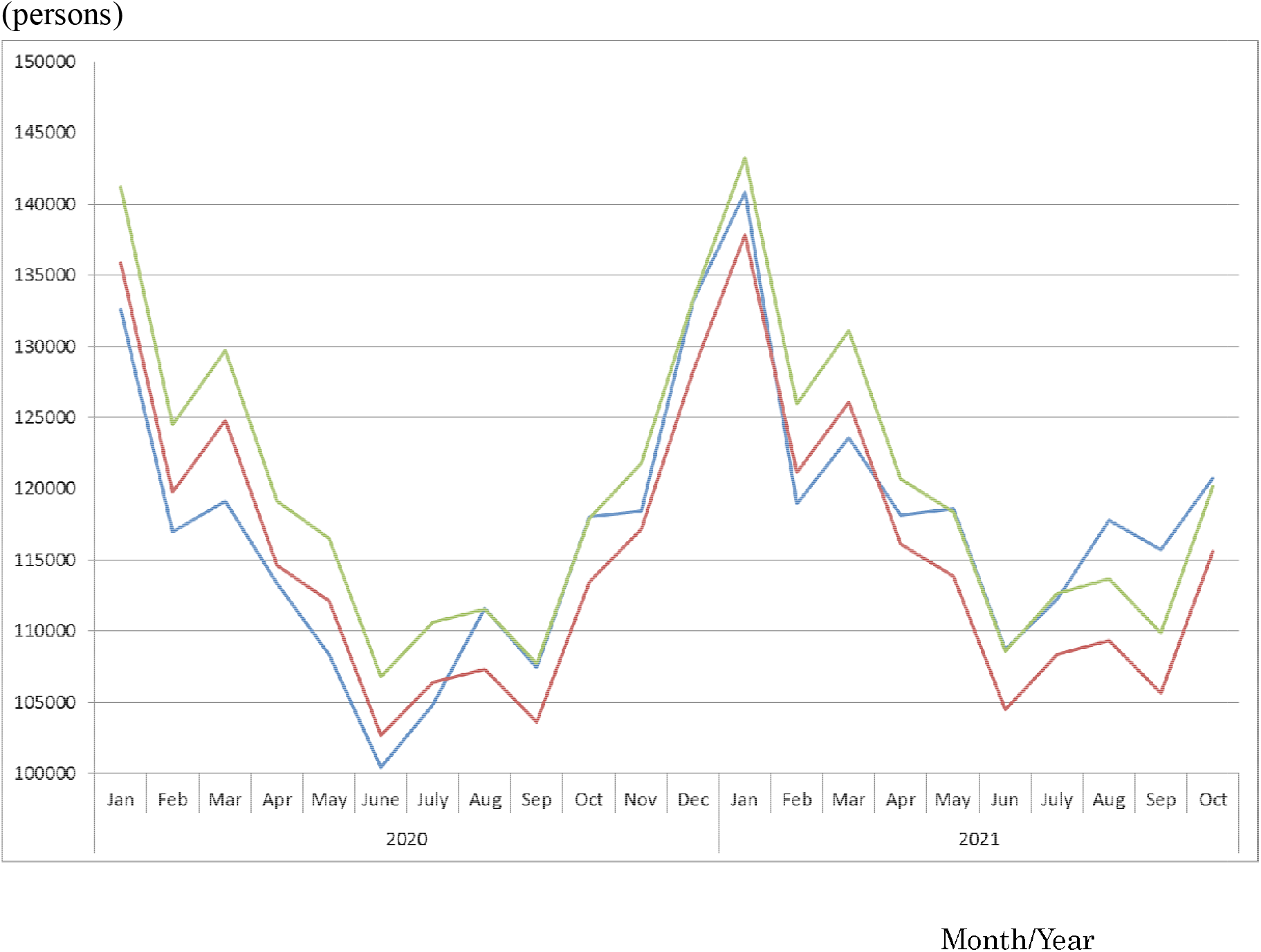
Observation of the estimated baseline and threshold since January 2020 in Japan Note: The blue line represents observations. The red line represents the estimated baseline. The green line shows its threshold.

Figure 3 and 4 showed the estimated result in Tokyo. We found 595 excess mortality in August and 150 excess mortality in September 76 in October, 458 in December, 2020, 44 in January, 60 in April, and 762 in August, 2021 which were 6.3, 1.7, 0.8, 4.1, 0.4, 0.6 and 8.0 % of the baseline.

**Figure 3:**
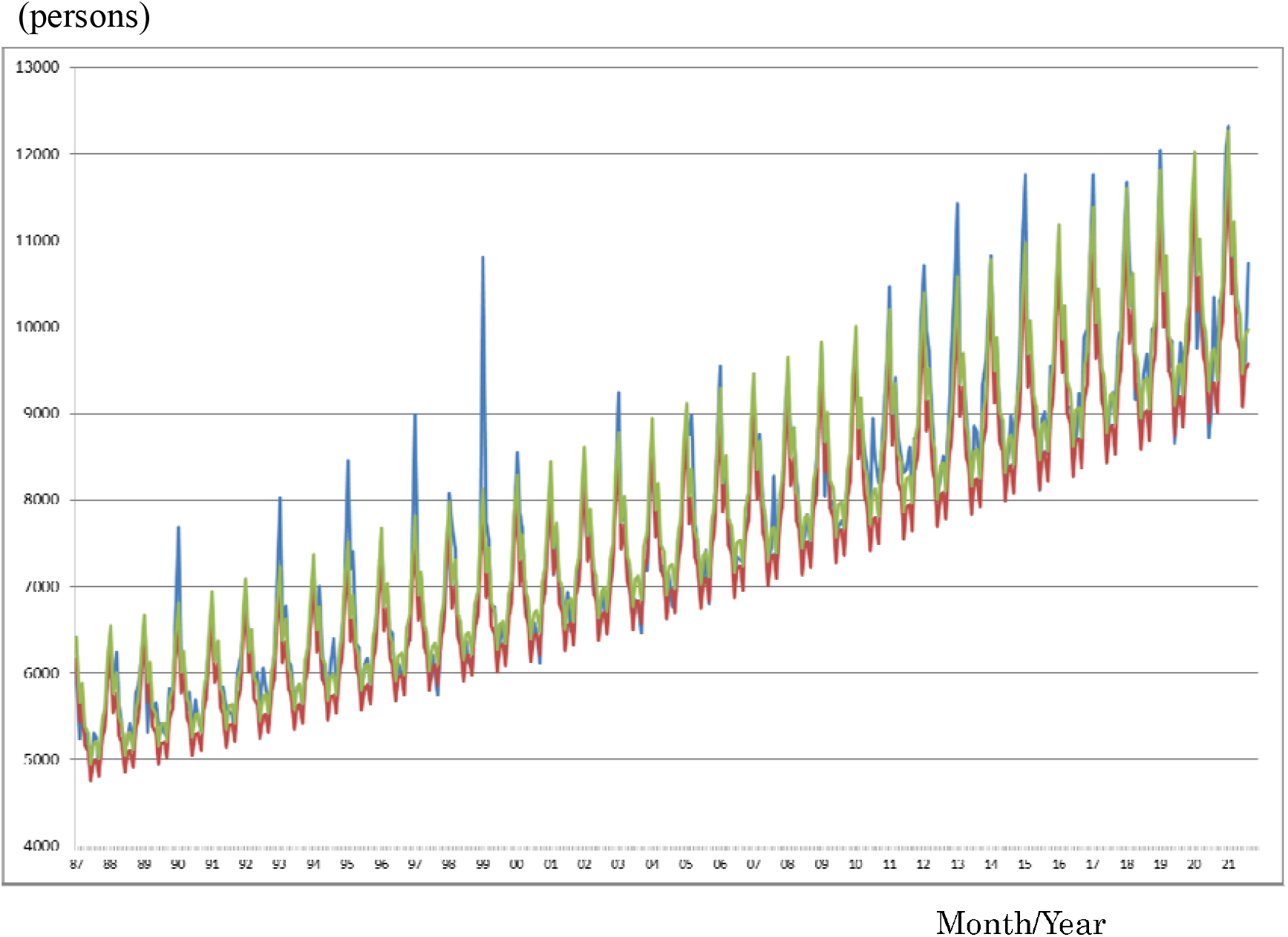
Observations of the estimated baseline and threshold since 1987 until August2021 in Tokyo Note: The blue line represents observations. The red line represents the estimated baseline. The green line shows its threshold.

**Figure 4:**
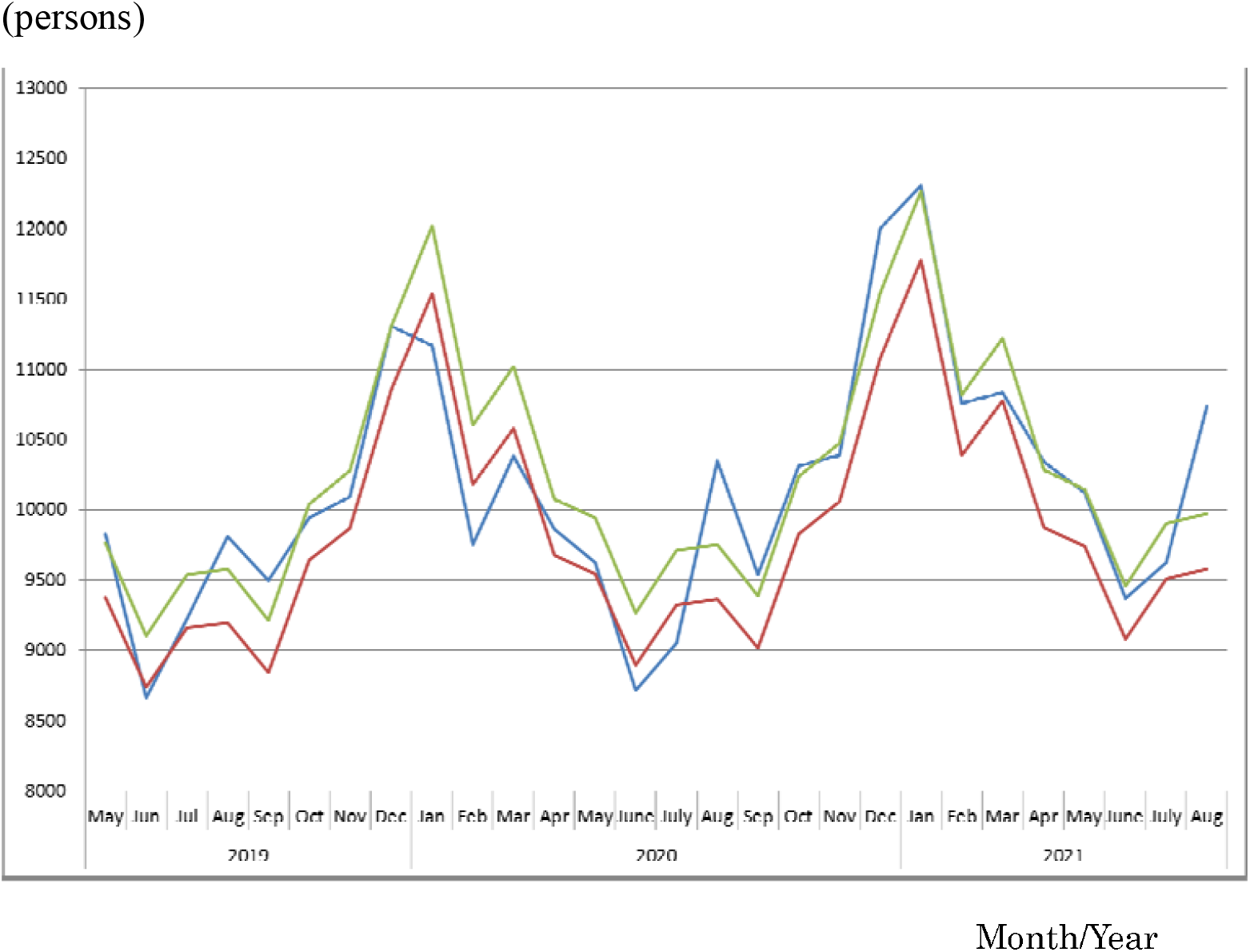
Observation of the estimated baseline and threshold since May 2019 until August 2021 in Tokyo Note: The blue line represents observations. The red line represents the estimated baseline. The green line shows its threshold.

## 4. Discussion

This study applied the NIID model to all causes of death to detect excess mortality attributable to COVID-19. We found huge number of excess mortality in August and September, 2021 up to about 10 thousands in the two months. in the whole of Japan. At the same time, only 2464 PCR-confirmed death were reported, which was less than quarter of excess mortality. Especially, in Tokyo, we found substantial excess mortality which was approximately 8% of the baseline at that time.

Some researchers in Japan have emphasized considerable excess mortality from all causes of death through June 2021 of around 49 thousand at maximum due to COVID-19 [20, 21] using the Farrington algorithm [22] and EuroMOMO [23], which was more than three times larger than the number of death confirmed by PCR test until July, 2021, 53 thousands. Their study measured excess mortalities as the gap between observation and beeline, not threshold as, in prefectures where observation was higher than threshold. Therefore, their estimated too huge excess mortality may seriously mislead the risk participation for COVID-19 among the general population. In particular, in Tokyo, they found 4575 excess mortality until June, 2021. It was more than twice higher than PCR confirmed death in Tokyo, 2211.

At the same time, they also found approximately 60.5 thousands negative excess mortality in Japan until June, 2021 and 3911 negative excess mortality in Tokyo until June, 2021. Conversely, we found only 3 persons as negative excess mortality in February, 2020 in Tokyo, and 751 negative excess mortality in April, 2020, in the whole of Japan, as shown in Figure 2 and 4. Such a huge negative excess mortality in their estimation may doubt validity of their procedures. In particular, they showed that some areas have excess mortality in a week but also have negative excess mortality in a few weeks later. How should we interpret this strange phenomena ? Because they did not provide any interpretation for these phenomena, they probably cannot understand and explain their results.

Their estimated baseline might be upward biased. It probably suggested that their adopted procedure have upper biased for excess mortality comparison with NIID model, which was suggested logically [24]. Moreover, they used only five years to estimate and thus volatility in data might be too small to obtain more appropriate threshold.

Using pneumonia death data instead of total death data might be better to evaluate excess mortality caused by COVID-19. However, application rule of the International Classification of Diseases was revised on January 2017, after which pneumonia deaths decreased by approximately 25%. April 2020 was the fourth April since that of 2017. However, excess mortality in pneumonia death should be our next challenge.

## 5. Conclusion

We found substantial excess mortality since the outbreak of COVID-19 had emerged in the whole of Japan in August and September, 2021, up to 10 thousands. We also substantial excess mortality in Tokyo in August, 2021, which corresponds to be 8% of the baseline at that time. It should be important to continue to monitor excess mortality of COVID-19 carefully hereafter.

The present study is based on the authors’ opinions: it does not reflect any stance or policy of their professionally affiliated bodies.

## Data Availability

Ministry of Health, Labour and Welfare. Preliminary statistics on demographics

https://www.mhlw.go.jp/toukei/list/81-1a.html

## 6. Acknowledgement

We acknowledge Dr. Nobuhiko Okabe, Kawasaki City Institute for Public Health, Dr.Kiyosu Taniguchi, National Hospital Organization Mie National Hospital, and Dr.Nahoko Shindo, WHO for their helpful support.

## 8. Ethical considerations

All information used for this study was published on the web site of MHLW [12]. Therefore, no ethical issue is presented.

